# Burden, Causation, and Particularities of long-COVID in African populations: A rapid systematic review

**DOI:** 10.1101/2023.01.13.23284305

**Authors:** Peter S. Nyasulu, Jacques L. Tamuzi, Rajiv T. Erasmus

## Abstract

**Background:** The global estimated prevalence of long COVID-19 is 43%, and the most common symptoms found globally are fatigue, confusion, or lack of confusion, and dyspnea, with prevalence rates of 23%, 14%, and 13%, respectively. However, long COVID still lacks an overall review in African populations. The aim of this review was to determine the prevalence of long COVID, its most common symptoms, comorbidities, and pathophysiological mechanisms.

**Methods:** A systematic review of long COVID in African populations was conducted. The random effects model was used to calculate the pooled prevalence rates (95% CI). If the results could not be pooled, a narrative synthesis was performed.

**Results:** We included 14 studies from 7 African countries, totaling 6,030 previously SARS-CoV-2 infected participants and 2,954 long COVID patients. Long COVID had a pooled prevalence of 41% [26%-56%]. Fatigue, dyspnea, and confusion or lack of concentration were the most common symptoms, with prevalence rates (95% CI) of 41% [26%-56%], 25% [12%-38%], and 40% [12%-68%], respectively. Long COVID was associated with advanced age, being female, more than three long COVID symptoms in the acute phase, initial fatigue and dyspnea, post-recovery stress, sadness, and sleep disturbances, and loss of appetite at symptoms onset, mild, moderate, and severe, pre-existing obesity, hypertension, diabetes mellitus, and the presence of any chronic illness (P _≤_0.05). According to our review, high micro clot and platelet poor plasma (PPP) viscosity explain the pathophysiology of long COVID.

**Conclusion:** Long COVID prevalence in Africa was comparable to the global prevalence. However, the prevalence of the most common symptoms was higher in Africa. Comorbidities associated with long COVID may lead to additional complications in African populations due to hypercoagulation and thrombosis.

## 1. Background

Post-Acute Sequelae of severe acute respiratory coronavirus 2 (SARS-CoV-2) (PASC), colloquially referred to as “long coronavirus disease (COVID)”, is a poorly understood condition characterized by prolonged COVID-19 symptoms and/or the development of new symptoms following the resolution of acute SARS-CoV-2 infection. While there is still no formal clinical definition of long COVID, findings reported in individuals experiencing long COVID include general symptoms (tiredness or fatigue that interferes with daily life, symptoms that get worse after physical or mental effort (also known as “post-exertional malaise”), fever, respiratory and heart symptoms (difficulty in breathing or shortness of breath, cough, chest pain, fast-beating or heart palpitations, neurological symptoms (difficulty thinking or concentrating, headache, insomnia, dizziness, pins-and-needles feelings, change in smell or taste, depression or anxiety), digestive symptoms (Gastroesophageal reflux disease (GERD), diarrhoea, and stomach pain) and other symptoms (hyperlipidaemia, thromboembolism, kidney disorders, joint or muscle pain, rash and changes in menstrual cycles).^1-4^ For example, National Institute for Health and Care Excellence (NICE) defined the “Long COVID” as “signs and symptoms that develop during/after the COVID-19 infection persisting for more than 4 weeks and could not be explained by any other diagnosis”. In this categorization, the long COVID consists of two categories, “ongoing symptomatic COVID-19_″_, which indicates the symptoms last for 4–12 weeks; and “post-COVID-19 syndrome”, which means symptom persistence beyond 12 weeks.^5,6^ Baig et al. suggested using the term “Chronic COVID syndrome (CCS)” as opposed to “Long-COVID” or “Long Haulers”.^7^ He also presented an organ-based staging of the illness to prioritize immediate care needs.^5,7^

As of 21 December 2022, Africa recorded approximately 9.4 million confirmed COVID-19 cases accounting for just 1.45 % of 649,2 million global COVID-19 confirmed cases^8^, despite containing 12.5% of the global population. ^9,10^ In Africa, the picture of long COVID is not well described as little research has been conducted in this emerging field. However, few studies have been conducted on the burden of long COVID in Africa, long COVID prevalence has been reported to vary across and within African countries. Dryden et al. reported the highest long COVID prevalence compared to other studies conducted worldwide. ^11^ However, a single study cannot reflect the prevalence of the African continent. While Peluso et al. concluded that HIV status strongly predicted the presence of long COVID. ^12^ Other studies found that diabetes, cardiovascular disease, and female sex were all linked to long COVID. ^13,14^ Given the high prevalence of these comorbidities on the African continent, an overall study focusing on African populations is required. We determined the burden of long COVID, prevalent symptoms, key findings, risk factors, and plausible pathophysiology in African populations in this review.

## 2. Methods

This section covers study selection strategy, study design, eligibility, inclusive and exclusive criteria, and quality of assessment and synthesis.

### 2.1 Study selection strategy

To ensure the reproducibility and transparency of our findings, studies were selected for systematic reviews (from January 2020 to December 2022) in accordance with the Preferred Reporting Items for Systematic Reviews and Meta-Analyses (PRISMA) guidelines.^15^ The studies were chosen based on the following key review research questions: How prevalent is long COVID in African populations? What are the most prevalent long COVID symptoms in African populations? What factors and comorbidities are associated with long COVID in African populations? To respond these questions, we searched PubMed, Medline, Google Scholar, the WHO COVID-19 Research Database, medRxiv, and bioRxiv for potential studies on long COVID-19 in African populations, employing the following MESH terms and key words as the search strategy: “Post-Acute COVID-19 Syndromes” OR “Long Haul COVID-19” OR “Long COVID” OR “Post-Acute Sequelae of SARS-CoV-2 Infection” OR “Post-COVID Conditions” OR “Post-COVID Condition” OR “Long-Haul COVID” AND “Comorbidities” AND “epidemiology” [Subheading] OR “Burden of Disease” AND “African populations” OR “African people” OR “Africans”. Only English-language studies were considered. To remove duplicates, studies were imported into the Endnote online tool. The authors took a step-by-step approach, beginning with systematic reviews. The remaining abstracts were screened by one reviewer, and all excluded abstracts were screened by another. One reviewer screened all full-text papers, and a second screened those that were not. When necessary, a third reviewer was brought in. One reviewer (JLT) extracted the data, and another (PSN) double-checked it. The study ID, setting, population, long COVID prevalence, comorbidities, and statistically significant findings were all included in the data. The Newcastle-Ottawa Scale (NOS) risk of bias assessment was used to evaluate the study’s quality.^16^

### 2.2 Study design eligible inclusion and exclusion criteria

We only included studies that looked at long COVID prevalence, symptoms, associated comorbidities, and other important findings in African populations. This review included people of all ages. We also included African studies on COVID symptoms that persisted. Furthermore, studies on asymptomatic COVID-19 and those assessing COVID recovery time were excluded from this review.

### 2.3 Data extraction and synthesis

We created a data extraction table to collect information related to the research questions. However, data were extracted for the four most common long COVID symptoms. Data extraction was completed by one reviewer (JLT) and independently checked by a second reviewer (PSN), with discrepancies resolved through discussion. Only evidence directly relevant to the review question was extracted. The data included in the meta-analysis, primarily long COVID prevalence and the four most common symptoms, were extracted into an Excel spreadsheet and the effect sizes (ES) and standard errors were calculated. Long COVID prevalence was calculated for each included study by dividing the total number of SARS-CoV-2 infected patients by the total number of long COVID. The number of COVID-19 patients with symptoms divided by the total number of long COVID patients yielded the prevalence for each long COVID symptom.

A meta-analysis was performed for long COVID prevalence and the four most common symptoms. According to the Cochrane handbook^17^, the prevalence rates were pooled using the random effects model because we expected significant heterogeneity (I^2^>50%). Subgroup analyses based on setting and long COVID were performed a priori. For small sample studies, Egger’s regression and Begg’s tests were used to assess publication bias. The P-value for statistical significance was set at 0.05 for all analyses. Stata (V.16, Stata Corp, College Station, Texas, USA) was used for the meta-analysis. Comorbidities and key findings were summarized in a narrative format.

## 3. Results

The databases yielded 453 articles, 39 duplicates were removed, and 414 titles and abstracts were screened for eligibility. 387 records were excluded because they were unsuitable for the review. Full texts were obtained for the 27 studies, but 13 were excluded because two were protocols, nine were reports, and two were letters to the editors. Then, 14 studies were included in our analysis, with 9 included in the meta-analysis and 5 included in the narrative synthesis. The review flow chart is shown in Figure 1.

**Figure 1:**
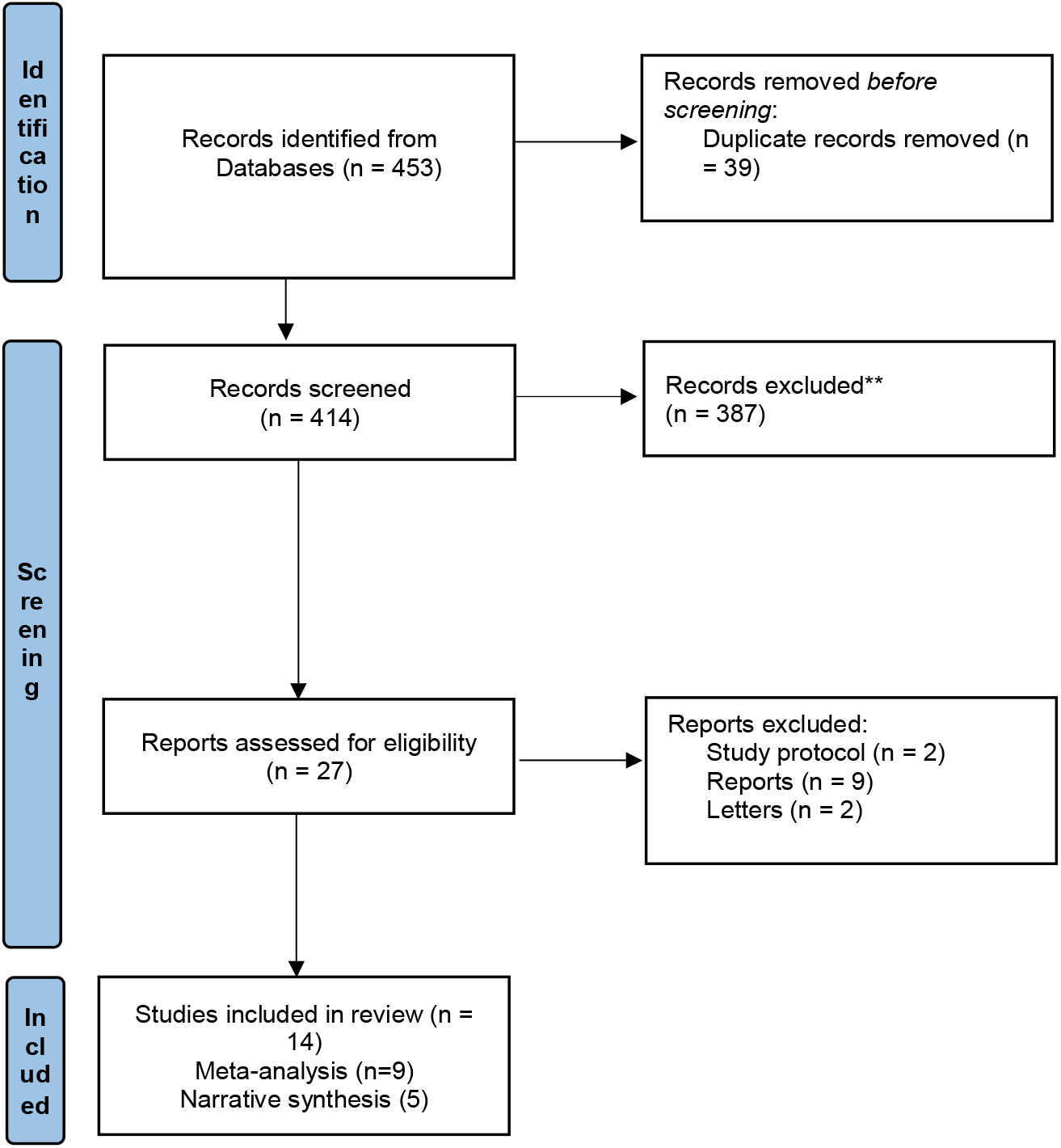
Flow chart of studies reviewing long COVID in African populations

### 3.1 Study and participant characteristics

Fourteen studies were included in this review. Among them, five were case control studies conducted in South Africa and Morocco^18-22^, three retrospective cohort studies undertaken in Nigeria and South Africa^23-25^, two prospective cohort studies (South Africa and Zambia)^11,26^, two retrospective cross-sectional studies done in South Africa and Egypt^27,28^ and two cross-Sectional studies undertaken Ghana and Egypt^29,30^. This review included a total of 6, 030 previously SARS-CoV-2 infected participants and 2,954 long COVID patients. Seven studies reported that long COVID was prevalent among females. ^11,18,20,25-29^ In contrast, three studies high prevalence rates among males.^23,24,30^ Long COVID range of age was 1 to 85 years.^11,18,19,22-24,27,28,30^ Methodological quality of the included studies was reported in Supplementary file 1.

### 3.2 Meta-analysis

The pooled results of long COVID prevalence (95% CI) in African populations revealed a 41% [26%-56%] overall prevalence (**Figure 2**). This prevalence, however, varied significantly across studies conducted in different African countries. Egypt had the highest prevalence, at 59% [55%-64%] (**Figure 2**). Nigeria had a prevalence rate of 49% [31%-66%]. (Figure 2). South Africa, Morocco, and Zambia had prevalence rates of 47% [7%-86%], 47% [38%-56%], and 15% [10%-20%], respectively (**Figure 2**). Ghana had the lowest prevalence of 2% [2%-3%] (**Figure 2**). The degree of heterogeneity between studies was high, with I^2^ = 99.5%.

**Figure 2:**
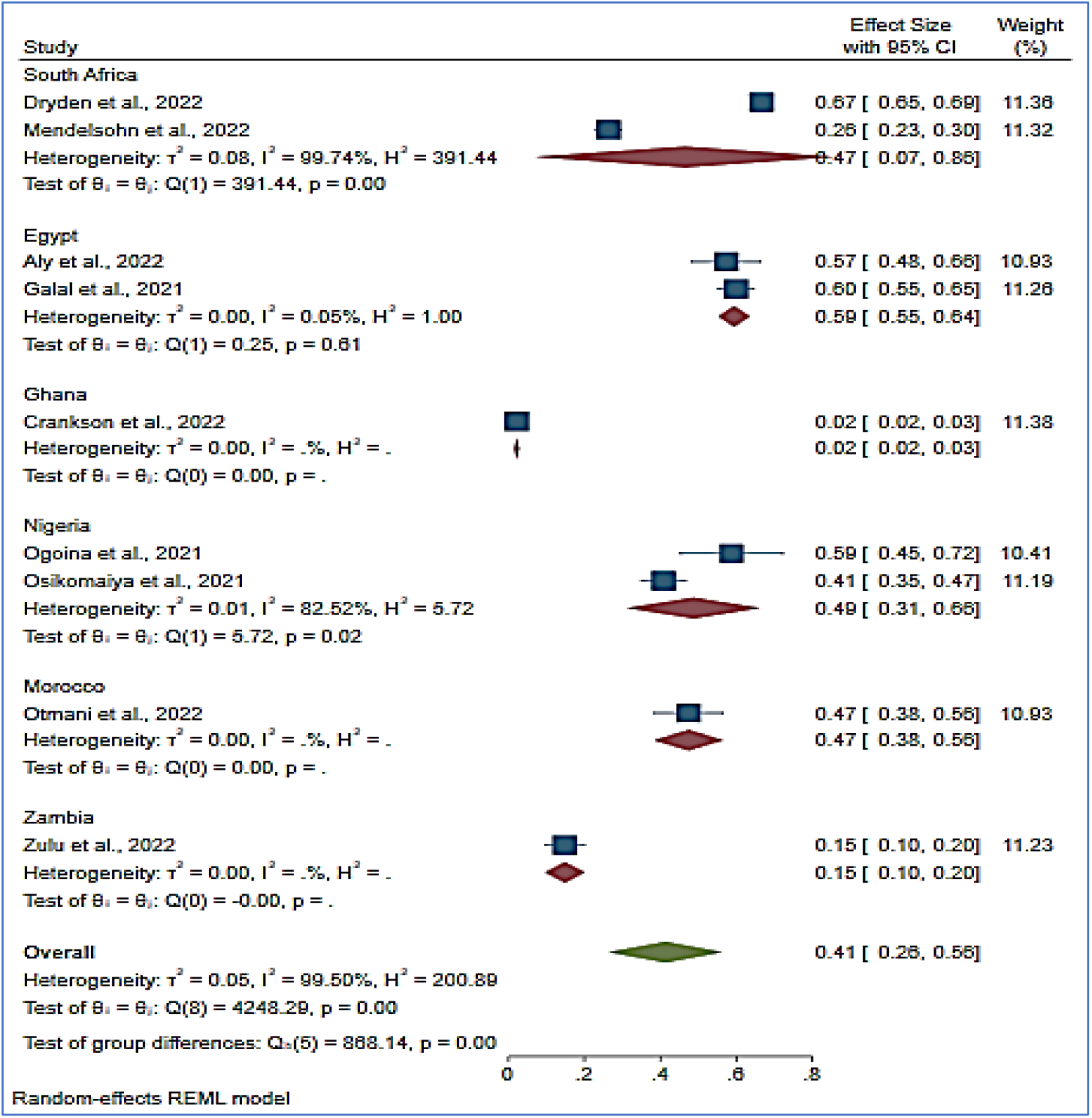
Forest plot of long COVID prevalence in African countries

The prevalence of the most common long COVID symptoms in African populations was assessed in **Figure 3**. According to the pooled prevalence, fatigue, dyspnea, and confusion or lack of concentration were the most common and prevalent long COVID symptoms among African populations. Fatigue, dyspnea, and confusion or lack of concentration were all prevalent (95% CI) at 41% [26%-56%], 25% [12%-38%], and 40% [12%-68%], respectively (**Figure 3**). A study of 845 long COVID participants found that fatigue, brain fog, loss of concentration and forgetfulness, dyspnea, and joint and muscle pains were the most common symptoms in African populations (**Table 1**). However, due to missing data, we were unable to include this in the meta-analysis.

**Figure 3:**
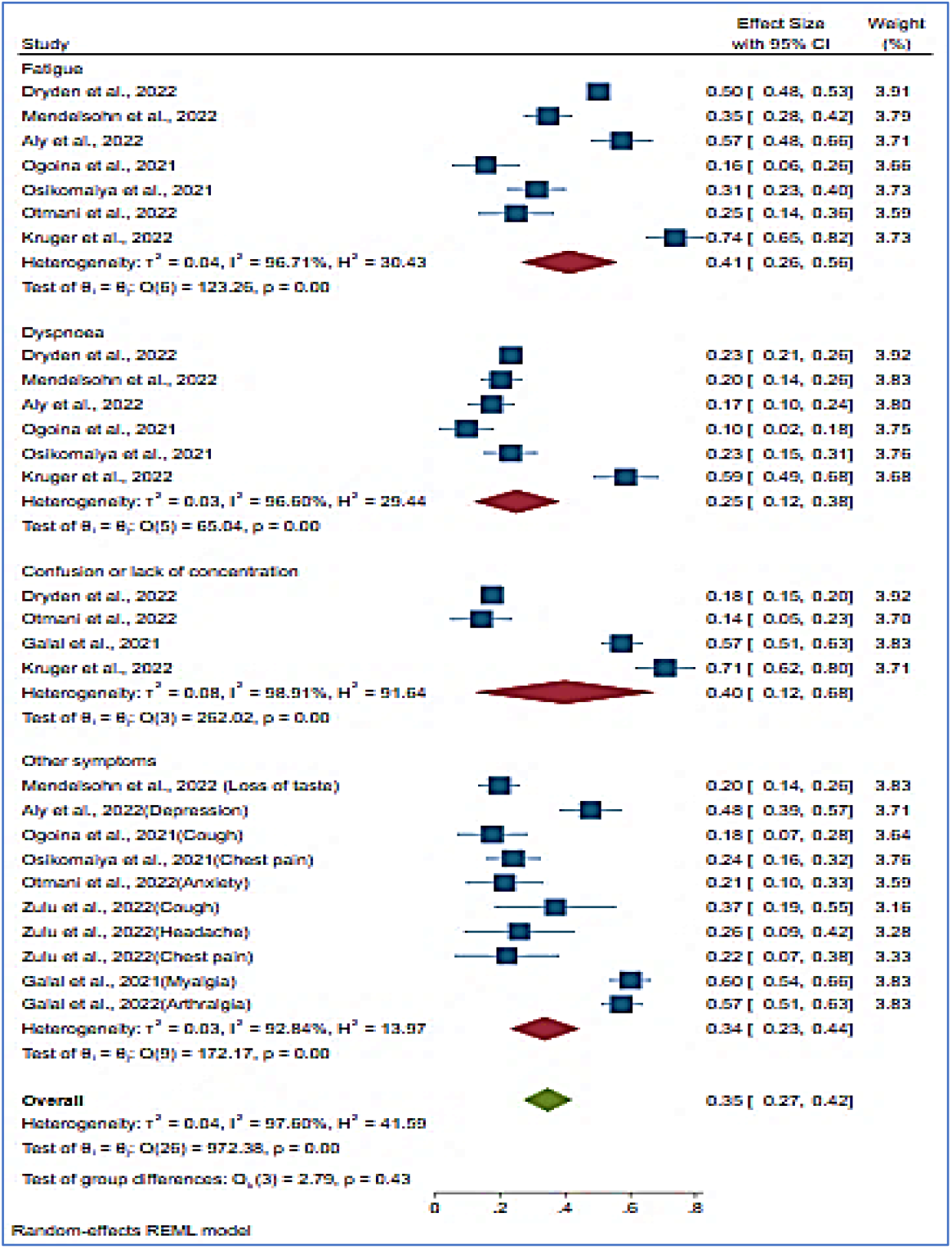
Forest plot of predominant symptoms of long COVID in African populations

**Table 1.**
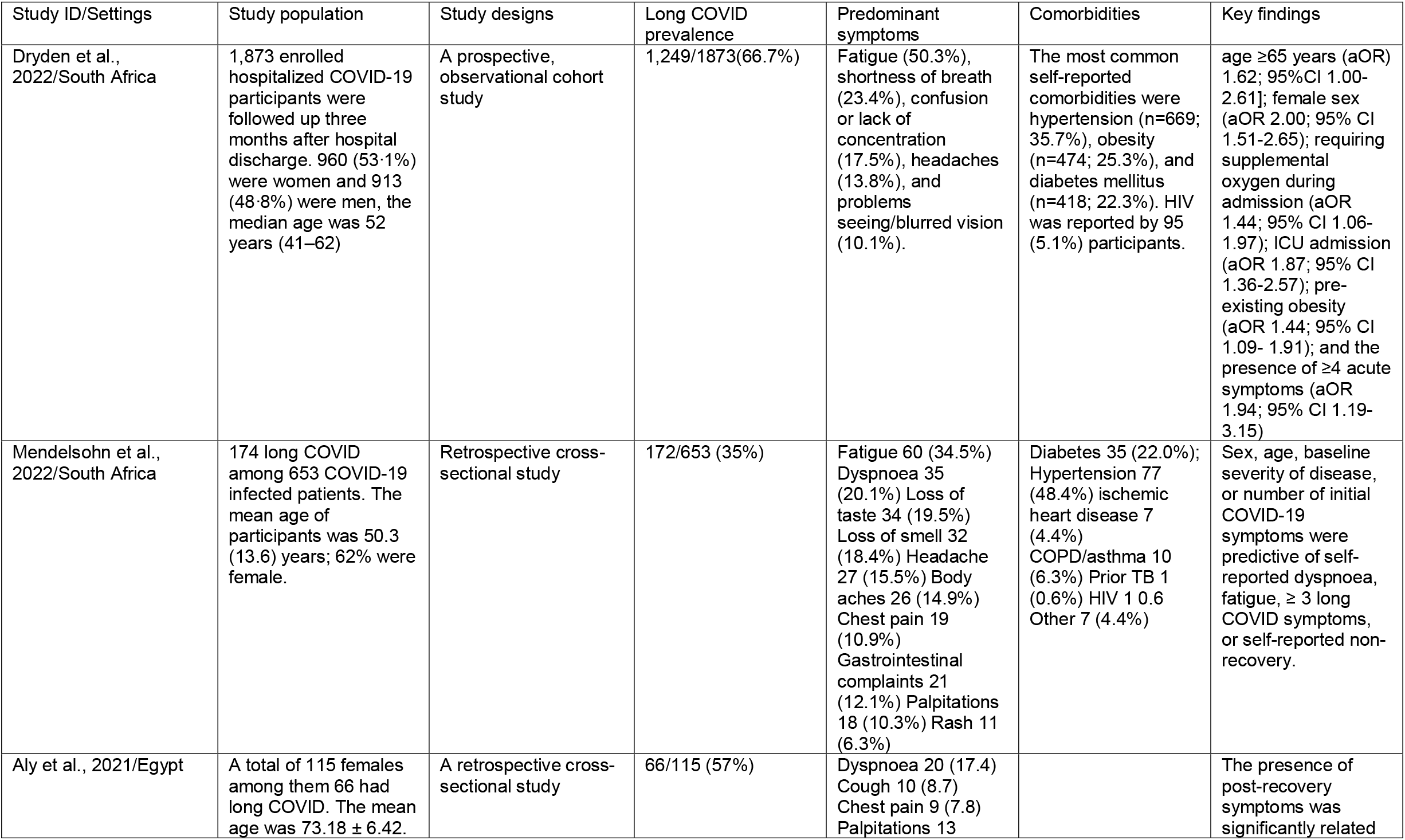
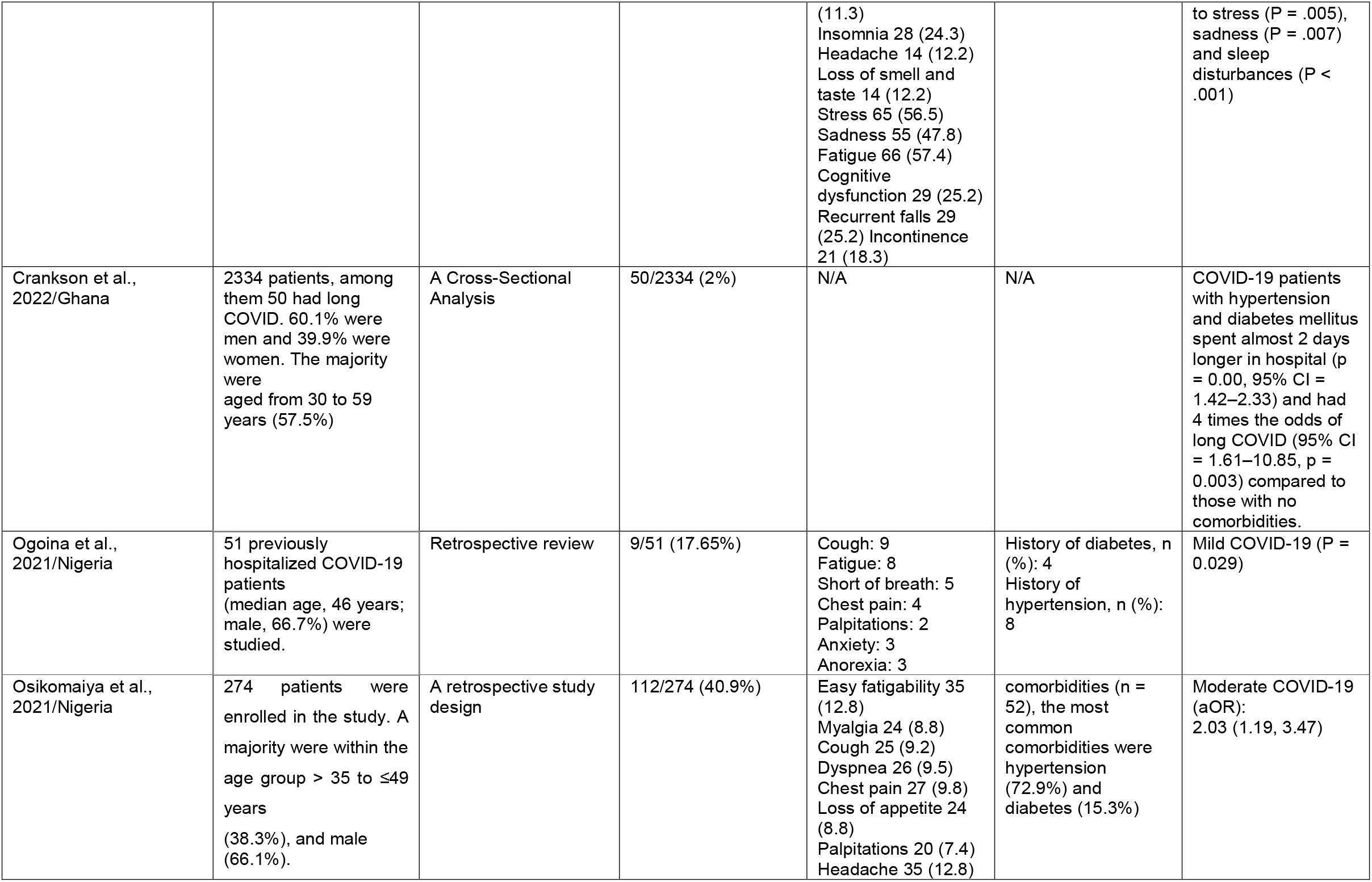
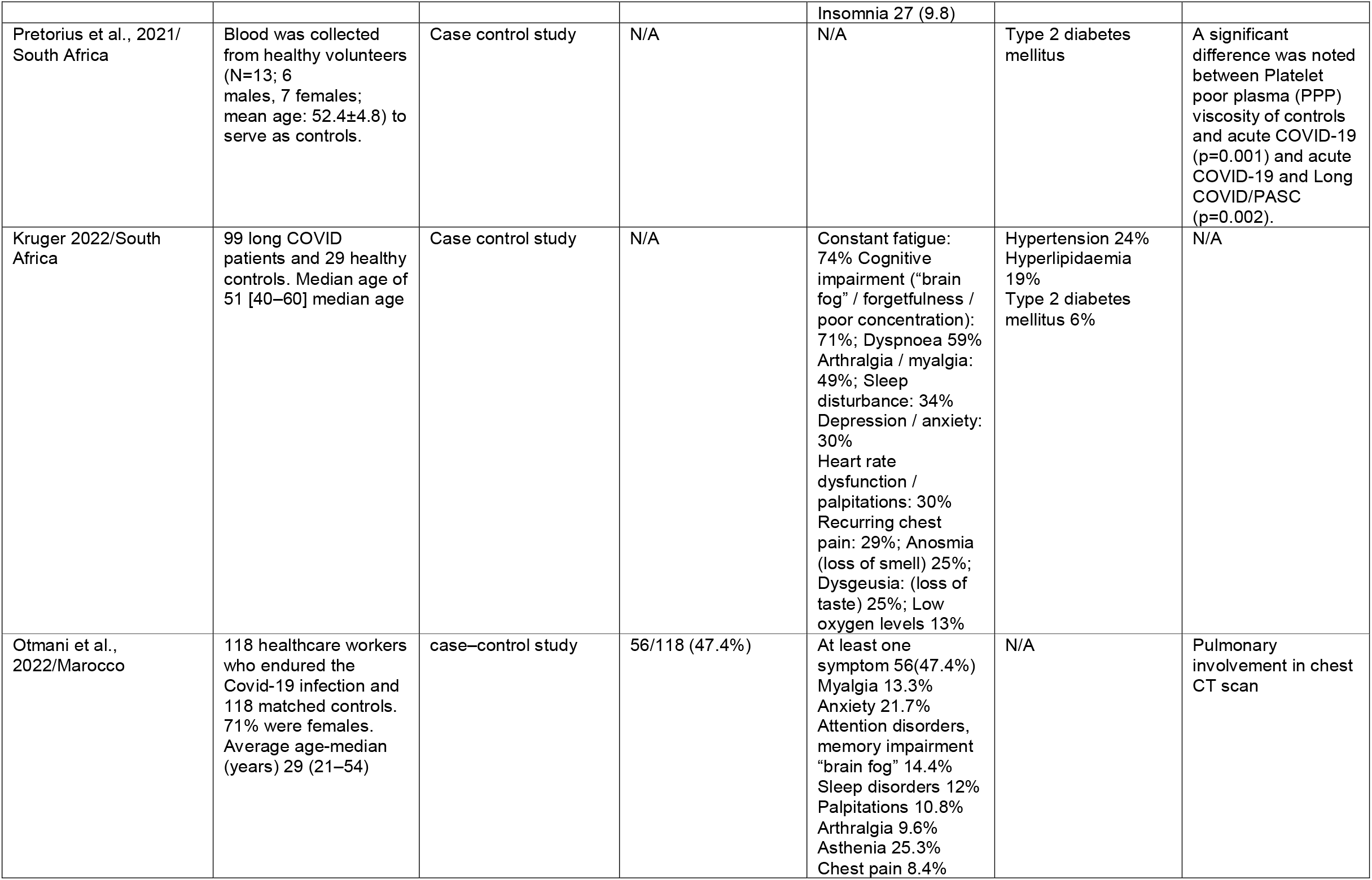
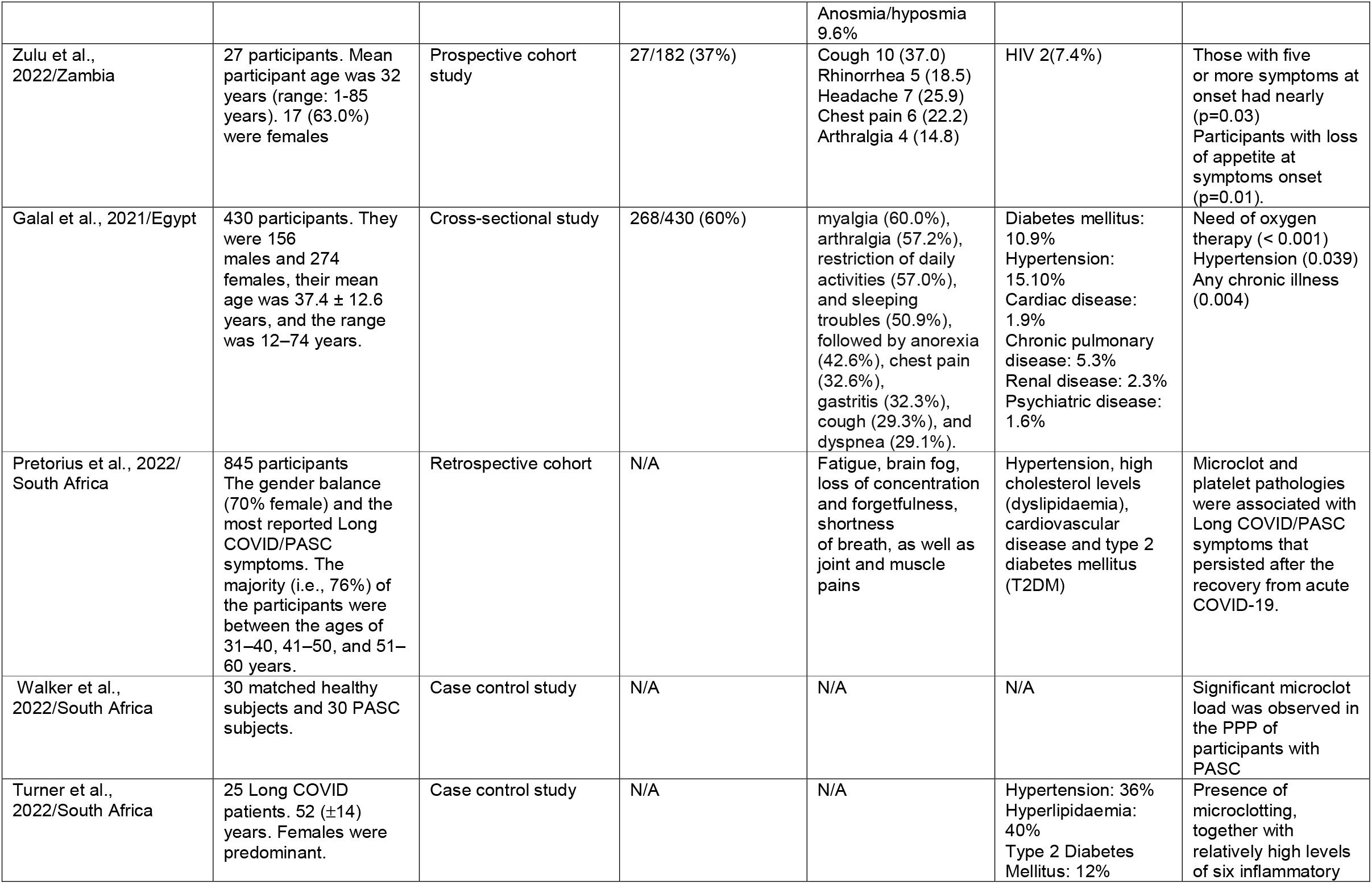
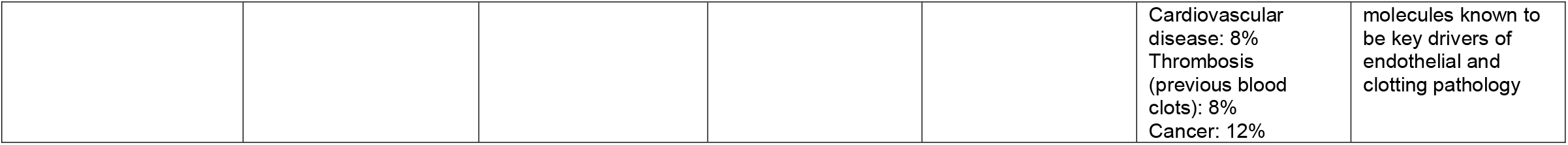
summarises the evidence-based studies of long COVID-19 in Africa

Other common symptoms were arthralgia, myalgia, depression, chest pain, headache, anxiety, and cough (**Figure 3**). Even though the test of heterogeneity between studies was high (I^2^ = 97.60%), the test of group difference was not statistically significant (P = 0.43).

Using Egger’s regression and Begg’s tests, we assessed the publication bias of nine studies that included long COVID prevalence in African populations. With z = 1.49 (P = 0.1538) and z = -0.52 (P = 1.3978), the publication bias was less likely.

### 3.3 Narrative synthesis

The narrative synthesis reported all statistically significant key findings (P 0.05) and important findings related comorbidities associated with long COVID and pathophysiology.

Long-COVID was statistically associated with advanced age and female sex in African populations (P _≤_0.05).^11,28^ In terms of clinical symptoms, long COVID was statistically associated with more than three long COVID symptoms in the acute phase, including initial fatigue and dyspnea, post-recovery stress, sadness and sleep disturbances, and loss of appetite at symptoms onset (P _≤_0.05).^11,26,27,28^ Our review also found that long COVID was statistically associated with mild, moderate, and severe COVID-19 (P _≤_0.05).^11,23,24,28^ Long COVID was also associated with supplemental oxygen during intensive care unit (ICU) admission and pulmonary involvement in chest CT scan (P _≤_0.05).^11,20,29^ Obesity, hypertension, diabetes mellitus, and the presence of any chronic illness were all statistically significant predictors of long COVID.^11,29,30^ The most common comorbidities found in long COVID in African populations were hypertension, obesity, hyperlipidaemia, diabetes mellitus, human immunodeficiency virus (HIV), ischemic heart disease, chronic obstructive pulmonary disease (COPD)/asthma, malignancies, previous tuberculosis (TB), renal disease, thrombosis (previous blood clots), and psychiatric diseases, as shown in **Table 1** and **Figure 4**. In terms of pathophysiology, African studies found that long COVID patients had higher microclot and platelet poor plasma (PPP) viscosity when compared to control groups.^18,21,22,25^ Furthermore, high levels of six inflammatory molecules were detected, including serum Amyloid A (SAA), -2 antiplasmin (−2AP), platelet factor 4 (PF4), Von Willebrand Factor (VWF), endothelial-leukocyte adhesion molecule 1 (E-selectin), and platelet endothelial cell adhesion molecule-1 (PECAM-1).^22^

**Figure 1.**
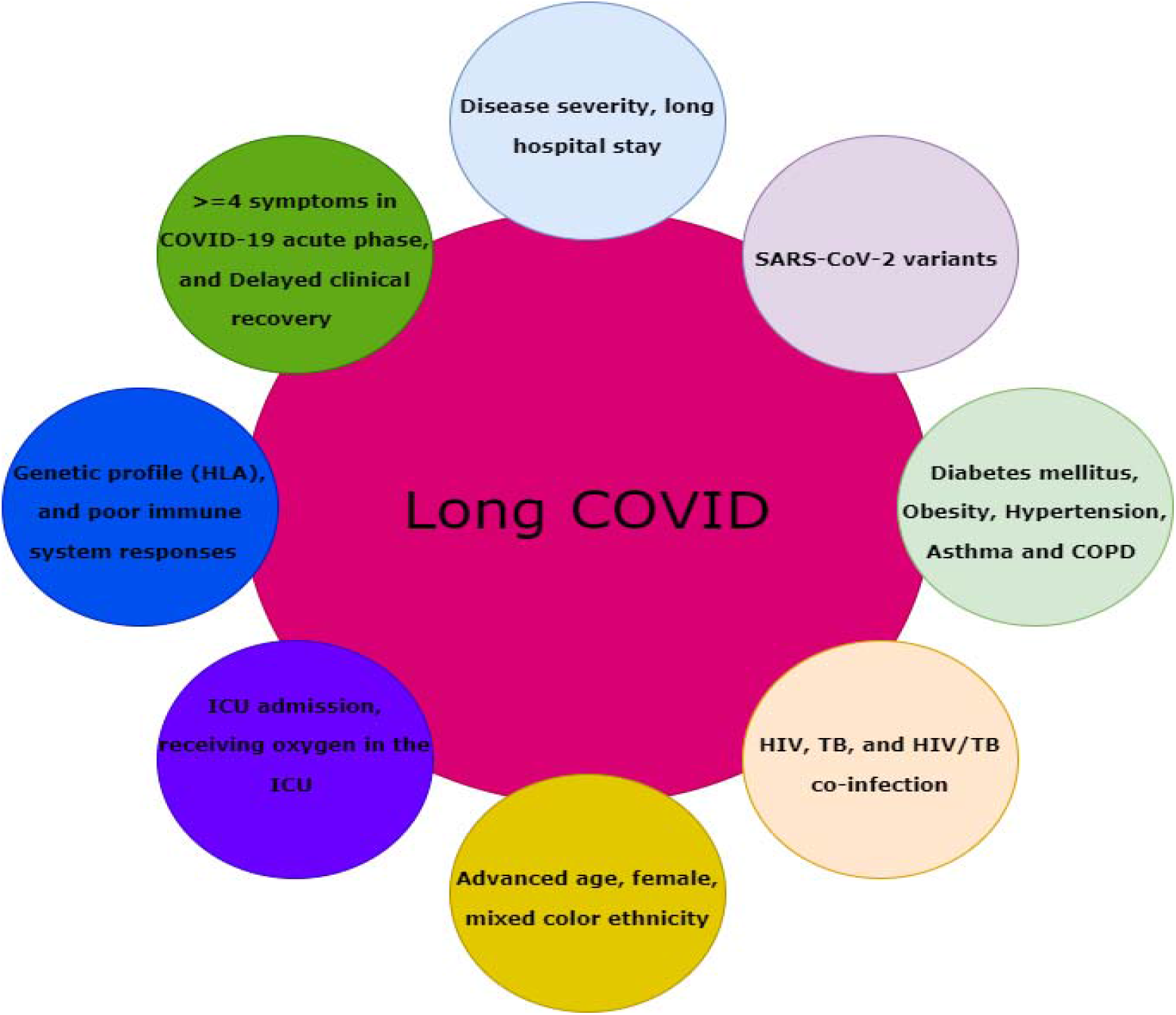
Potential long COVID risk factors in Africa.

## 4. Discussion

We conducted a systematic review to determine the burden of long COVID, prevalent symptoms, key findings, risk factors, and plausible pathophysiology in African populations. This review included fourteen studies on long COVID conducted in African populations. This study included 6,030 previously SARS-CoV-2 infected participants and 2,954 long COVID patients. Females outnumbered males among long COVID patients. The ages ranged from one to 85 years. The pooled prevalence (95% CI) of long COVID in the African population was 41% [26%-56%]. In comparison to a review of fifty included studies, 41 of which were meta-analysed, the global estimated pooled prevalence of long COVID-19 was 43% [39%-46%].^13^ Long COVID prevalence was higher in Africa than in North America [31% (21%-43%)], but lower in Europe [44% (32%-56%)] and Asia [51% (37%-65%)].^13^ The prevalence (95%CI) of the most common symptoms were 41% [26%-56%] fatigue, 40% [12%-68%] confusion or lack of concentration, and 25% [12%-38%] dyspnoea. Our result was also on same line with a global review showing that the fatigue was the most common symptom reported with a prevalence of 23% [17%-30%]^13,31^, followed by confusion or lack of concentration 14% [10%-19%]^13^ and dyspnoea 13% [11%-15%]^13^. Even though the African long COVID prevalence was slightly lower than the global prevalence, the Africans had higher rates of fatigue, confusion or lack of concentration, and dyspnea. Further research may be required to clarify these findings.

Our findings also revealed that Long-COVID was statistically associated with advanced age and being female in African populations.^11,28^ Our findings were consistent with the findings of Chen et al., who found that female sex was a risk factor for long COVID. Long COVID was statistically associated with more than three long COVID symptoms in the acute phase, including initial fatigue and dyspnea, post-recovery stress, sadness, and sleep disturbances, and loss of appetite at the onset of symptoms.^11,26-28^ Our findings support the results from previous systematic review and meta-analysis on long COVID symptoms.^13,31,32^

Our study also found that long COVID was statistically associated with mild, moderate, and severe COVID-19.^11,23,24,28^ This could be explained by a significant increase in the rates of thrombotic events following even a mild COVID-19 infection.^22,33^ Long COVID was also statistically associated with supplemental oxygen during ICU admission and pulmonary involvement in chest CT scan.^11,20,29^ A study that looked at the long-term outcomes of patients who needed ICU admission for severe COVID-19 found that the long-term effects of severe COVID-19 lasted more than six months.^34^ Similarly, a 12-month study found that a chest CT scan in acute COVID-19 correlates with pulmonary function and respiratory symptoms after SARS-CoV-2 infection.^35^ Obesity, hypertension, diabetes mellitus, and the presence of any chronic illness were all statistically significant predictors of long COVID.^11,29,30^ Preexisting conditions such as obesity and comorbidities were also found to be positively associated with post COVID-19 condition in multiple studies.^36,37^

In point of view pathophysiology, African studies demonstrated high microclot and Platelet poor plasma (PPP) viscosity were statistically higher in long COVID patients compared to the control groups.^18,21,22,25^ In addition, high levels of six inflammatory molecules SAA, _α_-2AP, PF4, VWF, E-selectin and PECAM-1, are known to be key drivers of endothelial and clotting pathology.^22^ In fact, long COVID was associated with additional distinctive innate and adaptive immune traits, consisting of a weaker initial inflammatory response.^38^ The viral tropism defined by the entry into cells through a widely expressed Angiotensin-Converting Enzyme 2 (ACE2) receptor makes it highly possible that many organs have the potential to undergo not only acute but also chronic damage, adding to the very diverse clinical picture of long COVID.^39,40^ Long COVID may generate further complications based on hypercoagulation and thrombosis associated to advanced age and pre-existing co-morbidities. As an example, in African populations, a study reported that two people living with HIV (PLWH) developed cardiovascular complications (myocardial infarction and arrhythmia) during the three months of follow-up.^41^ HIV status was strongly associated with long COVID, raising concerns that this condition might be common among PWH recovering from COVID-19.^12^ Certainly, two main hypotheses should be considered. Firstly, it is well known how COVID-19 may cause and worsen cardiovascular events, especially in patients who are at increased risk such as PLWH who also is in increased risk of hypertension, diabetes mellitus, dyslipidaemia, previous thrombosis and obesity.^42,43^ Secondly, in patients with known pre-existing cardiovascular risk factors, SAA, _α_-2AP, PF4, VWF, E-selectin and PECAM-1 induced by the presence of SARS-CoV-2 may have worsened and accelerated the cardiac damage and caused stress cardiomyopathy.^22,41,43,44^ Turner et al. explained long COVID as the interaction between endothelial dysfunction, chronic inflammation, and platelet activation generating the formation of micro clot, inducing the raised of the above mentioned six inflammatory molecules and hypercoagulation and thrombosis.^22^ This is significant because diabetes type 2 is a risk factor for long COVID in African populations and is associated with hypercoagulation states.^18^ Comorbidities associated with an increased risk of long COVID, and hypercoagulability include cardiovascular disease, renal failure, venous thrombosis, cancer, and COPD.^45^

Given that Africa has the lowest COVID-19 vaccination rate among continents, this could be another risk factor for long COVID in Africa. A systematic review of six studies (n=17,256,654 individuals) investigating the impact of vaccines before acute SARS-CoV-2 infection found a low level of evidence suggesting that vaccination before SARS-CoV-2 infection could reduce the risk of subsequent long-COVID infection.^46^ As with most common chronic conditions, the causation is not yet well understood. However, long COVID causation should be viewed as a multifactorial interaction between the individual co-morbidities, past medical history, SARS-CoV-2 variants, genetic factors, and ethnicity. **Figure 4** in our study has clearly highlighted the multifactorial factors of long COVID in African populations. The reason that some individuals are more prone to develop long COVID possibly lies in their genetic profile primarily related to the immune system, such as human leukocyte antigen (HLA).^5^ Long COVID may be the result of tissue/organ damage, or inflammatory and immune pathways dysfunction (including chronic inflammation, hypercoagulation, thrombosis, hyperactive immune cells, and autoimmunity because of molecular mimicry).^11,18,21,22,25^ This results to difference long COVID prevalence rates and symptoms among populations.

To the best of our knowledge, this is the first systematic review and meta-analysis of long COVID and its most common symptoms. This review will be useful in future research on long COVID and comorbidities in African populations, particularly comorbidities associated with thrombosis risk in African populations. The review’s main weaknesses were the small number of included studies, poor study design of included studies, missing data, and high heterogeneity between studies. However, publication bias was less likely because the Egger’s regression and Begg’s tests were not statistically significant with z = 1.49 (P = 0.1538) and z = -0.52 (P = 1.3978), respectively, and the test of group difference for long COVID symptoms was not statistically significant (P = 0.43).

## 5. Conclusion

This review looked at the various characteristics of long COVID in Africa. Despite having the lowest number of COVID-19 cases, the prevalence of long COVID cases in Africa was closer to the global prevalence. Even though fatigue, dyspnea, and confusion or lack of concentration were the most common symptoms globally and in Africa, their prevalence rates were higher in Africa. Long COVID was associated with advanced age, female sex, three long COVID symptoms in the acute phase, initial fatigue and dyspnea, post-recovery stress, sadness, and sleep disturbances, and loss of appetite at symptoms onset, mild, moderate, and severe, pre-existing obesity, hypertension, diabetes mellitus, and the presence of any chronic illness in African populations. According to our review, the pathophysiology of long COVID is explained by high microclot and PPP viscosity. This may raise the risk of additional cardiovascular complications in African populations. Future research should focus on long COVID, and comorbidities associated with hypercoagulation and thrombosis in African populations.

## Data Availability

All data produced in the present work are contained in the manuscript

## 6. Abbreviations

-2AP: -2 antiplasmin
ACE2: Angiotensin-Converting Enzyme 2
AIDS: acquired immune deficiency syndrome
ARDS: acute respiratory distress syndrome
BMI: body mass index
CDC: Centers for Disease Control and Prevention
CCS: Chronic COVID syndrome
CFS: chronic fatigue syndrome
COPD: Chronic obstructive pulmonary disease
COVID-19: Coronavirus disease 2019
E-selectin: endothelial-leukocyte adhesion molecule 1
GORD: Gastroesophageal reflux disease
HIV: human immunodeficiency virus
HLA: human leukocyte antigen
ICU: intensive care unit
MIS-A: multi-system inflammatory syndrome in adults
MIS: multisystem inflammatory syndrome
NICE: National Institute for Health and Care Excellence
PASC: Post-Acute Sequelae of SARS-CoV-2
PECAM-1: platelet endothelial cell adhesion molecule-1
PLWH: persons living with HIV
PF4: platelet factor 4
PPP: platelet poor plasma
RNA: ribonucleic acid
RT-PCR: Reverse transcription polymerase chain reaction
SAA: serum Amyloid A
SARS-CoV-2: Severe acute respiratory syndrome coronavirus 2
TB: tuberculosis
UN: United Nations
VWF: Von Willebrand Factor
WHO: World Health Organization

## 6. Footnotes

### Author Contributions

PSN conceived the review. PSN and JLT searched potential peer review manuscripts. PSN and JLT conducted the data extraction. JLT undertook the data analysis. PSN and JLT drafted the manuscript. PSN, JLT and RTE edited the manuscript. This version was reviewed and approved by all the authors.

### Informed Consent Statement

Not applicable

### Availability of data and materials

All Data and material are available in this manuscript.

### Ethical Approval statement

Not applicable

## Supplementary material 1: Quality assessment of included studies

**Table 2.**
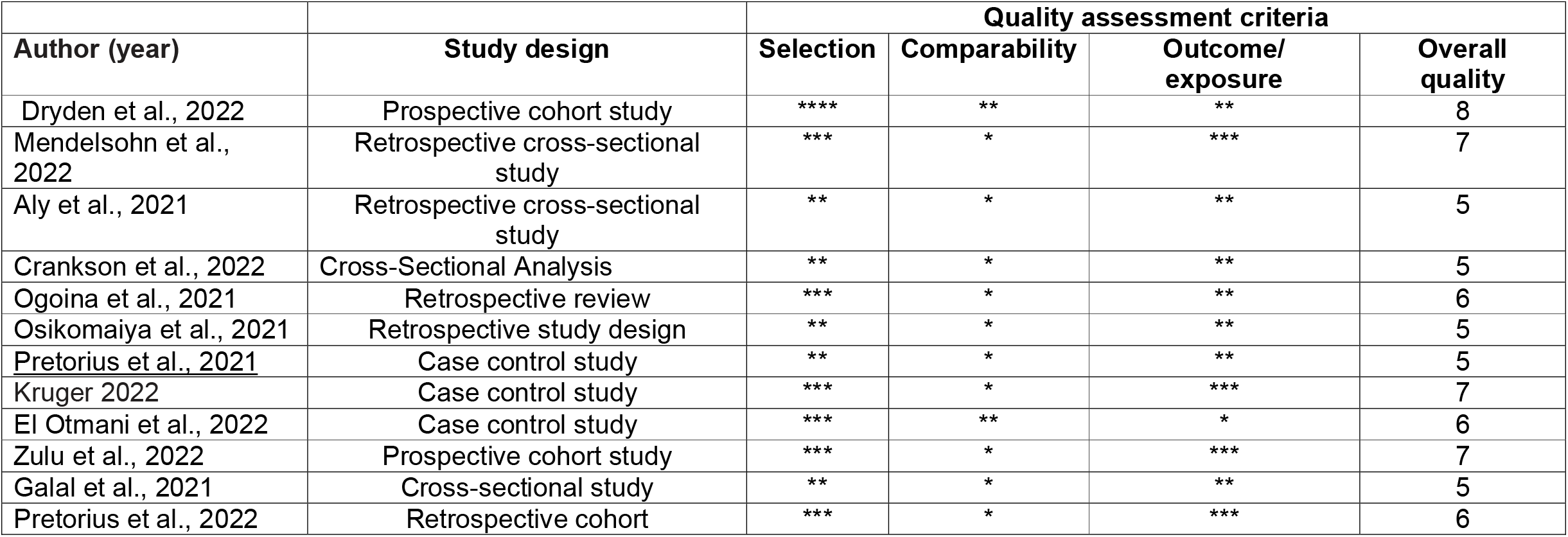
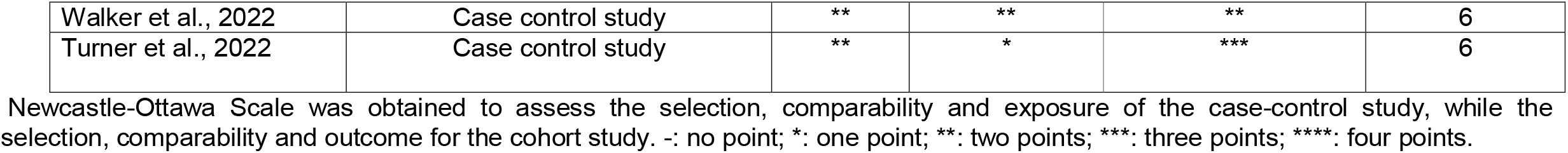
Quality assessment of included studies.

