## Supplemental Table 1 for "Burden, Causation, and Particularities of long-COVID in African populations: A rapid systematic review"

**Supplementary material 1: Quality assessment of included studies**

| Table 2. Quality assessment of included studies. | | | | | |
| --- | --- | --- | --- | --- | --- |
|  |  | **Quality assessment criteria** | | | |
| **Author (year)** | **Study design** | **Selection** | **Comparability** | **Outcome/ exposure** | **Overall quality** |
| Dryden et al., 2022 | Prospective cohort study | **** | ** | ** | 8 |
| Mendelsohn et al., 2022 | Retrospective cross-sectional study | *** | * | *** | 7 |
| Aly et al., 2021 | Retrospective cross-sectional study | ** | * | ** | 5 |
| Crankson et al., 2022 | Cross-Sectional Analysis | ** | * | ** | 5 |
| Ogoina et al., 2021 | Retrospective review | *** | * | ** | 6 |
| Osikomaiya et al., 2021 | Retrospective study design | ** | * | ** | 5 |
| Pretorius et al., 2021 | Case control study | ** | * | ** | 5 |
| Kruger 2022 | Case control study | *** | * | *** | 7 |
| El Otmani et al., 2022 | Case control study | *** | ** | * | 6 |
| Zulu et al., 2022 | Prospective cohort study | *** | * | *** | 7 |
| Galal et al., 2021 | Cross-sectional study | ** | * | ** | 5 |
| Pretorius et al., 2022 | Retrospective cohort | *** | * | *** | 6 |
| Walker et al., 2022 | Case control study | ** | ** | ** | 6 |
| Turner et al., 2022 | Case control study | ** | * | *** | 6 |

Newcastle-Ottawa Scale was obtained to assess the selection, comparability and exposure of the case-control study, while the selection, comparability and outcome for the cohort study. -: no point; *: one point; **: two points; ***: three points; ****: four points.
